# Predictors of Stroke Among U.S. Adults: A Survey-Weighted Analysis of the Behavioral Risk Factor Surveillance System, 2021–2023

**DOI:** 10.64898/2026.08.06.26359922

**Authors:** Karthik Samdeep Nayak, Abhay S Nirgude, Ranajit Das

## Abstract

**Background:** Stroke remains one of the leading causes of mortality, disability, and healthcare burden worldwide. Identifying demographic, socioeconomic, lifestyle, and clinical factors associated with stroke is essential for improving prevention strategies and reducing disease burden. This study aimed to identify independent predictors of stroke among U.S. adults using nationally representative Behavioral Risk Factor Surveillance System (BRFSS) data collected between 2021 and 2023.

**Methods:** A cross-sectional analysis was conducted using pooled BRFSS data from 2021–2023. Adults with complete information on stroke status and study variables were included in the multivariable analysis. Stroke status was determined from self-reported physician diagnosis. Survey-weighted multivariable logistic regression was performed to estimate adjusted odds ratios (aORs) and 95% confidence intervals (CIs) for demographic, socioeconomic, lifestyle, and clinical predictors while accounting for the complex BRFSS sampling design. Model discrimination was evaluated using receiver operating characteristic (ROC) curve analysis.

**Results:** Among 235,571 participants in the pooled dataset, stroke was more common among older adults and individuals with diabetes, poorer self-reported health, lower income, and smoking history. In the adjusted analysis, increasing age (aOR 1.04, 95% CI 1.04–1.04), diabetes (aOR 1.55, 95% CI 1.43– 1.67), current smoking (aOR 1.44, 95% CI 1.31–1.58), multiracial ethnicity (aOR 1.44, 95% CI 1.12– 1.82), Black race (aOR 1.31, 95% CI 1.15–1.50), and poorer general health (aOR 1.58, 95% CI 1.53– 1.64) were independently associated with higher odds of stroke. Conversely, Asian race (aOR 0.65, 95% CI 0.43–0.94), Hispanic ethnicity (aOR 0.65, 95% CI 0.54–0.77), higher income (aOR 0.92, 95% CI 0.90–0.93), and regular physical activity (aOR 0.86, 95% CI 0.80–0.92) were associated with lower odds of stroke. The final model demonstrated good discrimination, with an area under the ROC curve of 0.781 (95% CI 0.774–0.788).

**Conclusions:** Stroke among U.S. adults is independently associated with a combination of demographic, socioeconomic, lifestyle, and clinical factors. Diabetes, smoking, poor general health, and socioeconomic disadvantage remain important potentially modifiable contributors to stroke risk, whereas regular physical activity appears protective. These findings support targeted public health interventions focused on improving cardiometabolic health, promoting smoking cessation and physical activity, and addressing socioeconomic disparities to reduce the burden of stroke in the United States.

## Introduction

Stroke remains the second leading cause of death and one of the leading causes of long-term disability worldwide, imposing a substantial burden on individuals, healthcare systems, and society. According to the Global Burden of Disease (GBD) Study 2021, stroke continues to account for millions of deaths and disability-adjusted life years annually despite significant advances in prevention and acute care. The increasing prevalence of aging populations, metabolic disorders, and adverse lifestyle behaviors has further amplified the global burden of stroke, underscoring the need for effective strategies to identify individuals at high risk before the onset of clinical disease (1).

Stroke is a multifactorial disease resulting from complex interactions among demographic characteristics, socioeconomic status, lifestyle behaviors, and chronic medical conditions. Age is the strongest non-modifiable determinant of stroke risk, whereas diabetes mellitus, cigarette smoking, obesity, physical inactivity, and poor general health are among the most important modifiable contributors. Socioeconomic inequalities, including lower educational attainment and reduced household income, further influence stroke risk through disparities in healthcare access, health literacy, and management of chronic diseases. Consequently, accurate identification of these determinants is essential for developing effective prevention strategies and reducing stroke-related morbidity and mortality (1–3).

Several clinical prediction models have been developed to estimate an individual’s future risk of stroke. Among these, the Framingham Stroke Risk Profile, originally developed by Wolf et al. and subsequently updated by Dufouil et al., remains one of the most widely recognized tools for stroke risk assessment. However, these models primarily depend on clinical variables such as blood pressure measurements, antihypertensive treatment, atrial fibrillation, electrocardiographic findings, and other clinical assessments that may not be routinely available in large-scale public health surveys or community-based screening programs, thereby limiting their applicability in population-level epidemiological studies (2,3).

National health surveillance systems provide an opportunity to investigate stroke risk factors using large, representative populations. The Behavioral Risk Factor Surveillance System (BRFSS), administered annually by the U.S. Centers for Disease Control and Prevention (CDC), is the world’s largest continuously conducted health survey, collecting standardized information on demographic characteristics, health behaviors, chronic diseases, and preventive healthcare practices among U.S. adults. Owing to its nationally representative sampling framework and extensive sample size, the BRFSS has become an important resource for monitoring chronic diseases, evaluating health disparities, and informing public health policies (4).

Although numerous epidemiological studies have examined individual stroke risk factors, many have focused on selected clinical populations, regional cohorts, or a limited number of predictors. Furthermore, relatively few studies have comprehensively evaluated demographic, socioeconomic, lifestyle, and clinical determinants of stroke using recent nationally representative BRFSS data while accounting for the complex survey sampling design. Such analyses are essential for generating robust population-level evidence that accurately reflects stroke risk across diverse demographic groups and supports the development of targeted prevention strategies.

Therefore, the present study aimed to identify independent predictors of self-reported physician-diagnosed stroke among U.S. adults using pooled BRFSS data collected between 2021 and 2023. Using survey-weighted multivariable logistic regression, we evaluated the associations between demographic characteristics, socioeconomic status, lifestyle behaviors, and clinical factors with stroke while accounting for the complex sampling design of the BRFSS. We further assessed the discriminative performance of the final multivariable model using receiver operating characteristic (ROC) curve analysis.

## Materials and Methods

### Study Design and Data Source

This cross-sectional study utilized publicly available data from the Behavioral Risk Factor Surveillance System (BRFSS) collected during the 2021–2023 survey cycles. The BRFSS is an annual, state-based health surveillance program administered by the U.S. Centers for Disease Control and Prevention (CDC) that collects information on health-related risk behaviors, chronic health conditions, preventive healthcare utilization, and demographic characteristics among non-institutionalized adults aged 18 years or older residing in the United States. The BRFSS employs a complex, multistage probability sampling design incorporating stratification, clustering, and sampling weights to generate nationally representative population estimates. Data are collected through standardized telephone interviews using both landline and cellular telephone sampling frames. The BRFSS methodology has been described previously (4).

The three annual BRFSS datasets (2021, 2022, and 2023) were harmonized into a single analytical dataset following standard preprocessing procedures. After data cleaning, duplicate removal, variable harmonization, and quality assessment, the combined dataset consisted of 235,571 adult participants, of whom 7,127 (3.0%) reported a history of physician-diagnosed stroke.

### Study Population

Participants aged 18 years or older with available information on stroke status were eligible for inclusion. Individuals with missing outcome data were excluded from the analysis. Variables exhibiting extensive structural missingness, including hypertension and heavy alcohol consumption, were excluded from multivariable modelling because missing values were primarily attributable to differences in questionnaire administration across survey years rather than random missingness. Similarly, previous myocardial infarction and previous coronary heart disease variables were excluded because these variables contained no meaningful variation in the final analytical dataset after data preprocessing. Complete-case analysis was performed for the multivariable regression model, consistent with the default behavior of generalized linear modelling in R v4.5.2.

This study involved secondary analysis of publicly available, de-identified BRFSS data obtained from the U.S. Centers for Disease Control and Prevention. Because no identifiable participant information was used, additional institutional ethical approval and informed consent were not required. The study was conducted in accordance with the principles outlined in the Declaration of Helsinki.

### Outcome Variable

The primary outcome was self-reported physician-diagnosed stroke. Participants were classified as having experienced stroke if they responded affirmatively to the BRFSS question asking whether a healthcare professional had ever informed them that they had suffered a stroke. The outcome variable was analyzed as a binary variable (stroke: yes/no).

### Predictor Variables

Potential predictors were selected based on previous epidemiological evidence and biological plausibility. Variables included:

- **Demographic variables**: age (continuous), sex (male/female), and race/ethnicity (American Indian/Alaska Native, Asian, Black, Hispanic, Multiracial, Native Hawaiian/Pacific Islander, Other, and White).
- **Anthropometric variable**: body mass index (BMI, kg/m^2^).
- **Clinical variable**: self-reported diabetes mellitus.
- **Lifestyle variables**: smoking status (never, former, current) and physical activity (active/inactive).
- **Socioeconomic variables**: educational attainment and household income.
- **Self-perceived health**: general health status measured using the BRFSS five-category Likert scale ranging from excellent to poor.

Continuous variables were retained in their original form, whereas categorical variables were analyzed using clinically appropriate reference categories.

History of coronary heart disease and myocardial infarction were excluded as covariates because these variables exhibited no variation in the final analytical dataset following cohort selection. Hypertension (Structurally missing for all 2022 participants) and heavy alcohol consumption (Structurally missing for 2022–2023 survey cycles) were not included in the primary multivariable analysis because of substantial structural missingness across pooled BRFSS survey cycles, which would have resulted in the exclusion of an entire survey year from the complete-case analysis.

### Data Quality Assessment

Prior to statistical analysis, extensive data quality assessment was performed. Duplicate observations were identified and removed where appropriate. Variable distributions, data types, missing values, and coding consistency were examined across survey years. Missingness patterns were evaluated graphically and quantitatively to distinguish random from structurally missing variables. Variables demonstrating substantial structural missingness due to survey design differences between study years were excluded from inferential analyses rather than being imputed. Continuous variables were examined for implausible values and summary statistics were inspected to ensure consistency across survey cycles.

### Statistical Analysis

Continuous variables are presented as mean ± standard deviation (SD), whereas categorical variables are summarized as frequencies and percentages. Differences between participants with and without stroke were evaluated using the Wilcoxon rank-sum test for continuous variables and the Pearson chi-square test for categorical variables.

Univariable logistic regression analyses were initially performed to examine the association between each predictor and stroke. Variables considered clinically relevant were subsequently included in the multivariable logistic regression model irrespective of their univariable statistical significance. Adjusted odds ratios (aORs) and corresponding 95% confidence intervals (CIs) were estimated.

Because the BRFSS employs a complex sampling design, the primary analysis used survey-weighted multivariable logistic regression, incorporating the BRFSS sampling weights, primary sampling units (PSUs), and stratification variables to obtain nationally representative effect estimates. Survey weights were applied using the survey package in R to account for unequal probabilities of selection and non-response.

Multicollinearity among predictors was assessed using Variance Inflation Factors (VIFs), with values below 5 considered indicative of acceptable collinearity. Model discrimination was evaluated using receiver operating characteristic (ROC) curve analysis, and predictive performance was quantified by the area under the ROC curve (AUC) with corresponding 95% confidence intervals. Adjusted odds ratios were additionally visualized using a forest plot displaying effect estimates and confidence intervals.

All statistical analyses were performed using R version 4.5.2 (R Foundation for Statistical Computing, Vienna, Austria). Data manipulation and visualization were conducted using the tidyverse, ggplot2, broom, gtsummary, pROC, car, survey, and related R packages. All statistical tests were two-sided, and a P value <0.05 was considered statistically significant.

## Results

### Participant Characteristics

A total of 235,571 participants from the pooled 2021–2023 Behavioral Risk Factor Surveillance System (BRFSS) dataset were included in the descriptive analysis, of whom 7,127 (3.0%) reported a history of physician-diagnosed stroke and 228,444 (97.0%) reported no history of stroke. The mean age of the study population was 54 ± 18 years, whereas participants with stroke were substantially older than those without stroke (66 ± 14 vs. 54 ± 18 years, P < 0.001). The missingness in the data is shown in Figure 1.

**Figure 1:**
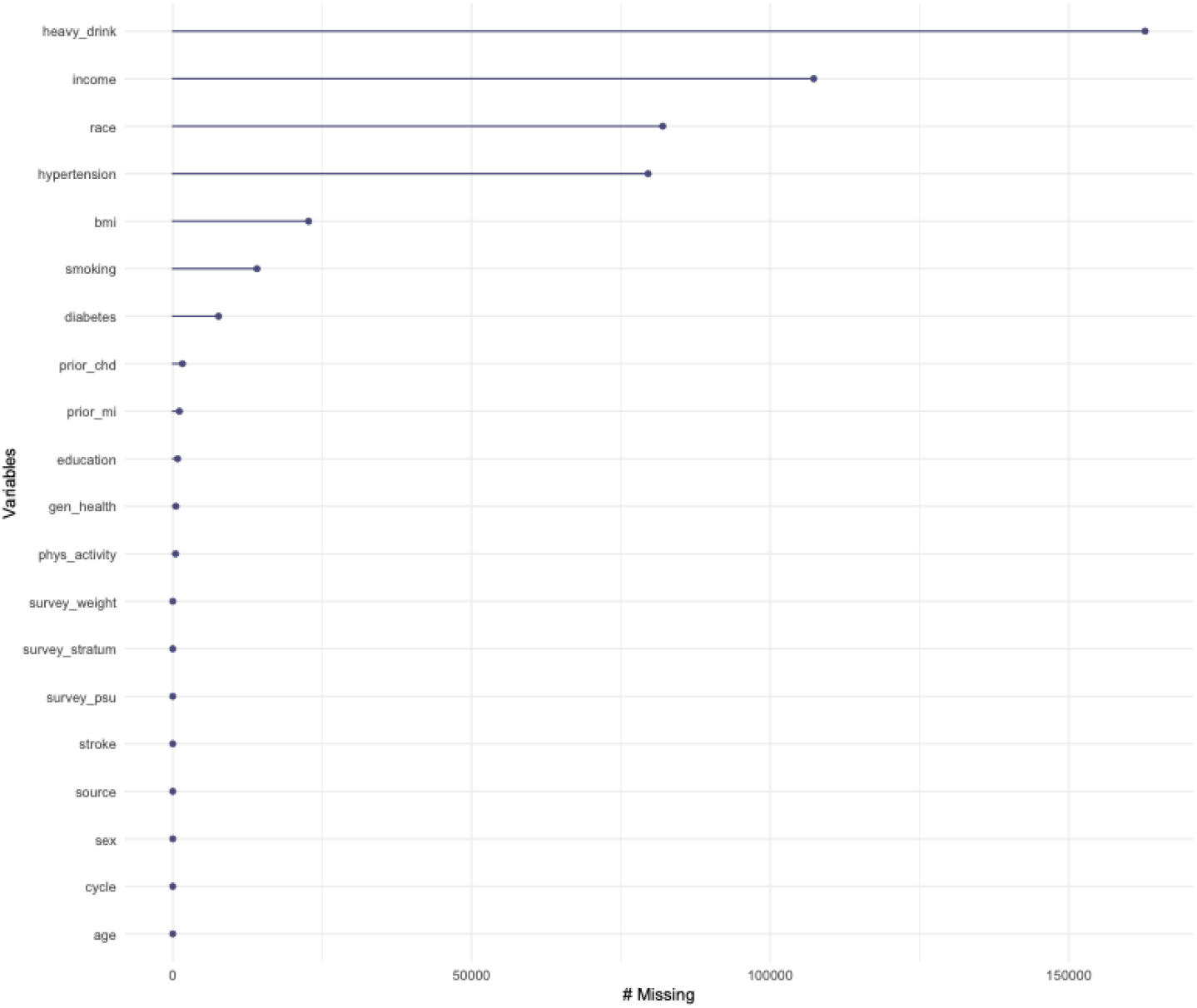
Missingness in the BRFSS data by various categories

Significant differences were observed between participants with and without stroke across most demographic, socioeconomic, lifestyle, and clinical characteristics (Table 1). Individuals with stroke were more likely to have diabetes (28% vs. 12%), report current or former smoking, be physically inactive, have lower household income, and report poorer general health compared with participants without stroke (all P < 0.001). Participants with stroke also had a slightly higher mean body mass index than those without stroke (29 ± 7 vs. 28 ± 7 kg/m^2^, P < 0.001). Sex and racial distributions also differed significantly between groups.

**Table 1:** Demographic. Socioeconomic, and Clinical characteristics of the participants.

| Characteristic | Overall<br>N = 235,571<sup>1</sup> | No Stroke<br>N = 228,444<sup>1</sup> | Stroke<br>N = 7,127<sup>1</sup> | p-value<sup>2</sup> |
| --- | --- | --- | --- | --- |
| age | 54 ± 18 | 54 ± 18 | 66 ± 14 | <0.001 |
| sex |  |  |  | <0.001 |
| female | 128,115 (54%) | 123,997 (54%) | 4,118 (58%) |  |
| male | 107,456 (46%) | 104,447 (46%) | 3,009 (42%) |  |
| race |  |  |  | <0.001 |
|  | 82,011 (35%) | 79,520 (35%) | 2,491 (35%) |  |
| aian | 2,405 (1.0%) | 2,304 (1.0%) | 101 (1.4%) |  |
| asian | 4,344 (1.8%) | 4,286 (1.9%) | 58 (0.8%) |  |
| black | 11,994 (5.1%) | 11,493 (5.0%) | 501 (7.0%) |  |
| hispanic | 15,507 (6.6%) | 15,237 (6.7%) | 270 (3.8%) |  |
| multiracial | 3,553 (1.5%) | 3,431 (1.5%) | 122 (1.7%) |  |
| nhpi | 752 (0.3%) | 730 (0.3%) | 22 (0.3%) |  |
| other | 1,323 (0.6%) | 1,280 (0.6%) | 43 (0.6%) |  |
| white | 113,682 (48%) | 110,163 (48%) | 3,519 (49%) |  |
| bmi | 28 ± 7 | 28 ± 7 | 29 ± 7 | <0.001 |
| Unknown | 22,742 | 22,219 | 523 |  |
| diabetes | 27,646 (12%) | 25,716 (12%) | 1,930 (28%) | <0.001 |
| Unknown | 7,663 | 7,360 | 303 |  |
| smoking |  |  |  | <0.001 |
| Never | 136,942 (62%) | 133,592 (62%) | 3,350 (50%) |  |
| Former | 58,179 (26%) | 55,955 (26%) | 2,224 (33%) |  |
| Current | 26,358 (12%) | 25,183 (12%) | 1,175 (17%) |  |
| Unknown | 14,092 | 13,714 | 378 |  |
| phys_activity |  |  |  | <0.001 |
| Inactive | 54,707 (23%) | 51,955 (23%) | 2,752 (39%) |  |
| Active | 180,387 (77%) | 176,037 (77%) | 4,350 (61%) |  |
| Unknown | 477 | 452 | 25 |  |
| education |  |  |  | <0.001 |
| 1 | 351 (0.1%) | 343 (0.2%) | 8 (0.1%) |  |
| 2 | 4,448 (1.9%) | 4,254 (1.9%) | 194 (2.7%) |  |
| 3 | 8,483 (3.6%) | 8,034 (3.5%) | 449 (6.3%) |  |
| 4 | 57,835 (25%) | 55,717 (24%) | 2,118 (30%) |  |
| 5 | 63,384 (27%) | 61,268 (27%) | 2,116 (30%) |  |
| 6 | 100,262 (43%) | 98,040 (43%) | 2,222 (31%) |  |
| Unknown | 808 | 788 | 20 |  |
| income |  |  |  | <0.001 |
| 1 | 5,371 (4.2%) | 5,070 (4.1%) | 301 (7.1%) |  |
| 2 | 5,412 (4.2%) | 5,060 (4.1%) | 352 (8.3%) |  |
| 3 | 7,159 (5.6%) | 6,765 (5.5%) | 394 (9.3%) |  |
| 4 | 10,219 (8.0%) | 9,674 (7.8%) | 545 (13%) |  |
| 5 | 21,992 (17%) | 21,033 (17%) | 959 (23%) |  |
| 6 | 25,277 (20%) | 24,457 (20%) | 820 (19%) |  |
| 8 | 26,936 (21%) | 26,396 (21%) | 540 (13%) |  |
| 10 | 12,660 (9.9%) | 12,475 (10%) | 185 (4.4%) |  |
| 11 | 13,263 (10%) | 13,113 (11%) | 150 (3.5%) |  |
| Unknown | 107,282 | 104,401 | 2,881 |  |
| gen_health |  |  |  | <0.001 |
| 1 | 40,703 (17%) | 40,352 (18%) | 351 (5.0%) |  |
| 2 | 82,221 (35%) | 80,738 (35%) | 1,483 (21%) |  |
| 3 | 75,652 (32%) | 73,190 (32%) | 2,462 (35%) |  |
| 4 | 28,555 (12%) | 26,702 (12%) | 1,853 (26%) |  |
| 5 | 7,914 (3.4%) | 6,977 (3.1%) | 937 (13%) |  |
| Unknown | 526 | 485 | 41 |  |

<sup>1</sup> Mean ± SD; n (%)
<sup>2</sup> Wilcoxon rank sum test; Pearson's Chi-squared test

### Univariable Logistic Regression Analysis

Univariable logistic regression analysis demonstrated significant associations between stroke and several demographic, socioeconomic, lifestyle, and clinical variables (Table 2). Increasing age, diabetes mellitus, current smoking, former smoking, lower income, physical inactivity, poorer general health, and several racial/ethnic groups were significantly associated with stroke before adjustment. These findings supported their inclusion in the multivariable regression model based on both statistical significance and clinical relevance.

**Table 2:**
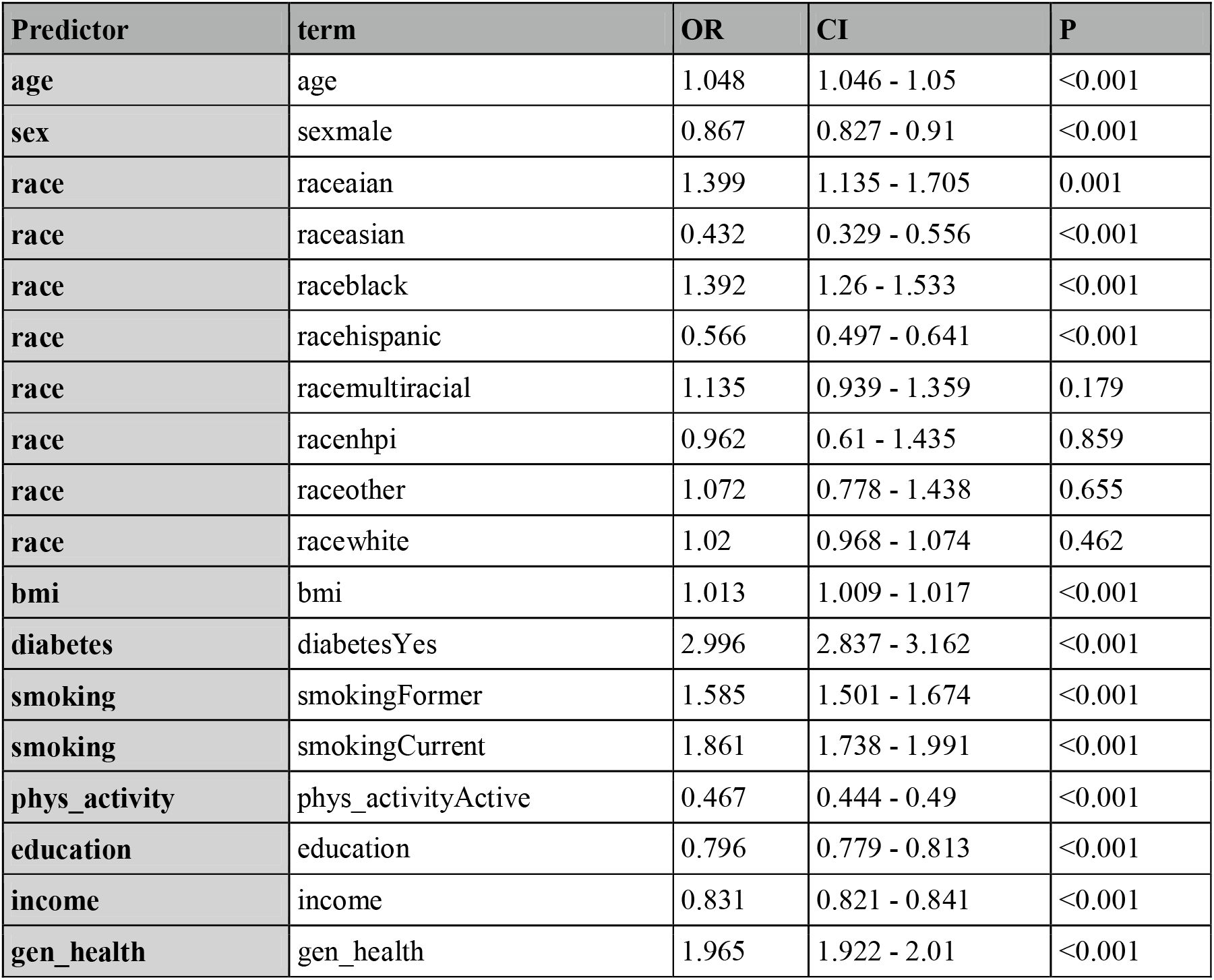
Univariate Logistic Regression.

### Multivariable Logistic Regression Analysis

After adjustment for potential confounding variables, several predictors remained independently associated with stroke (Table 3).

**Table 3:** Multivariate Logistic Regression.

| Characteristic | OR | 95% CI | p-value |
| --- | --- | --- | --- |
| age | 1.04 | 1.04, 1.04 | <0.001 |
| sex |  |  |  |
| female | — | — |  |
| male | 1.04 | 0.98, 1.12 | 0.2 |
| race |  |  |  |
| race | — | — |  |
| aian | 1.29 | 0.98, 1.67 | 0.061 |
| asian | 0.65 | 0.43, 0.94 | 0.031 |
| black | 1.31 | 1.15, 1.50 | <0.001 |
| hispanic | 0.65 | 0.54, 0.77 | <0.001 |
| multiracial | 1.44 | 1.12, 1.82 | 0.003 |
| nhpi | 1.20 | 0.66, 2.00 | 0.5 |
| other | 1.00 | 0.62, 1.53 | >0.9 |
| white | 0.96 | 0.89, 1.04 | 0.3 |
| bmi | 1.00 | 0.99, 1.00 | 0.2 |
| diabetes |  |  |  |
| No | — | — |  |
| Yes | 1.55 | 1.43, 1.67 | <0.001 |
| smoking |  |  |  |
| Never | — | — |  |
| Former | 1.12 | 1.03, 1.21 | 0.005 |
| Current | 1.44 | 1.31, 1.58 | <0.001 |
| phys_activity |  |  |  |
| Inactive | — | — |  |
| Active | 0.86 | 0.80, 0.92 | <0.001 |
| education | 1.01 | 0.98, 1.05 | 0.4 |
| income | 0.92 | 0.90, 0.93 | <0.001 |
| gen_health | 1.58 | 1.53, 1.64 | <0.001 |

Abbreviations: CI = Confidence Interval, OR = Odds Ratio

Increasing age remained one of the strongest independent predictors, with each additional year associated with a 4% increase in the odds of stroke (aOR = 1.04, 95% CI: 1.04–1.04, P < 0.001). Participants with diabetes had 55% higher odds of stroke than those without diabetes (aOR = 1.55, 95% CI: 1.43–1.67, P < 0.001). Similarly, current smokers (aOR = 1.44, 95% CI: 1.31–1.58, P < and former smokers (aOR = 1.12, 95% CI: 1.03–1.21, P = 0.005) exhibited significantly greater odds of stroke compared with never smokers. Poorer self-rated general health was also independently associated with increased stroke risk (aOR = 1.58, 95% CI: 1.53–1.64, P < 0.001).

Conversely, regular physical activity was associated with significantly reduced odds of stroke (aOR = 0.86, 95% CI: 0.80–0.92, P < 0.001). Higher household income also demonstrated a protective association (aOR = 0.92, 95% CI: 0.90–0.93, P < 0.001). Among racial and ethnic groups, Asian participants (aOR = 0.65, 95% CI: 0.43–0.94, P = 0.031) and Hispanic participants (aOR = 0.65, 95% CI: 0.54–0.77, P < 0.001) had significantly lower odds of stroke, whereas Black participants (aOR = 1.31, 95% CI: 1.15–1.50, P < 0.001) and multiracial participants (aOR = 1.44, 95% CI: 1.12–1.82, P = 0.003) had significantly higher odds. Sex, body mass index, education level, Native Hawaiian/Pacific Islander ethnicity, White race, Other race, and American Indian/Alaska Native ethnicity were not independently associated with stroke after adjustment. Multicollinearity was assessed using variance inflation factors (VIFs). All predictors had VIF values below 1.4, indicating negligible multicollinearity.

### Model Performance

The multivariable logistic regression model demonstrated good discriminative performance, achieving an area under the receiver operating characteristic curve (AUC) of 0.781 (95% CI: 0.774–0.788) (Figure 2). The ROC curve indicated satisfactory discrimination between participants with and without stroke, supporting the predictive performance of the final multivariable model.

**Figure 2:**
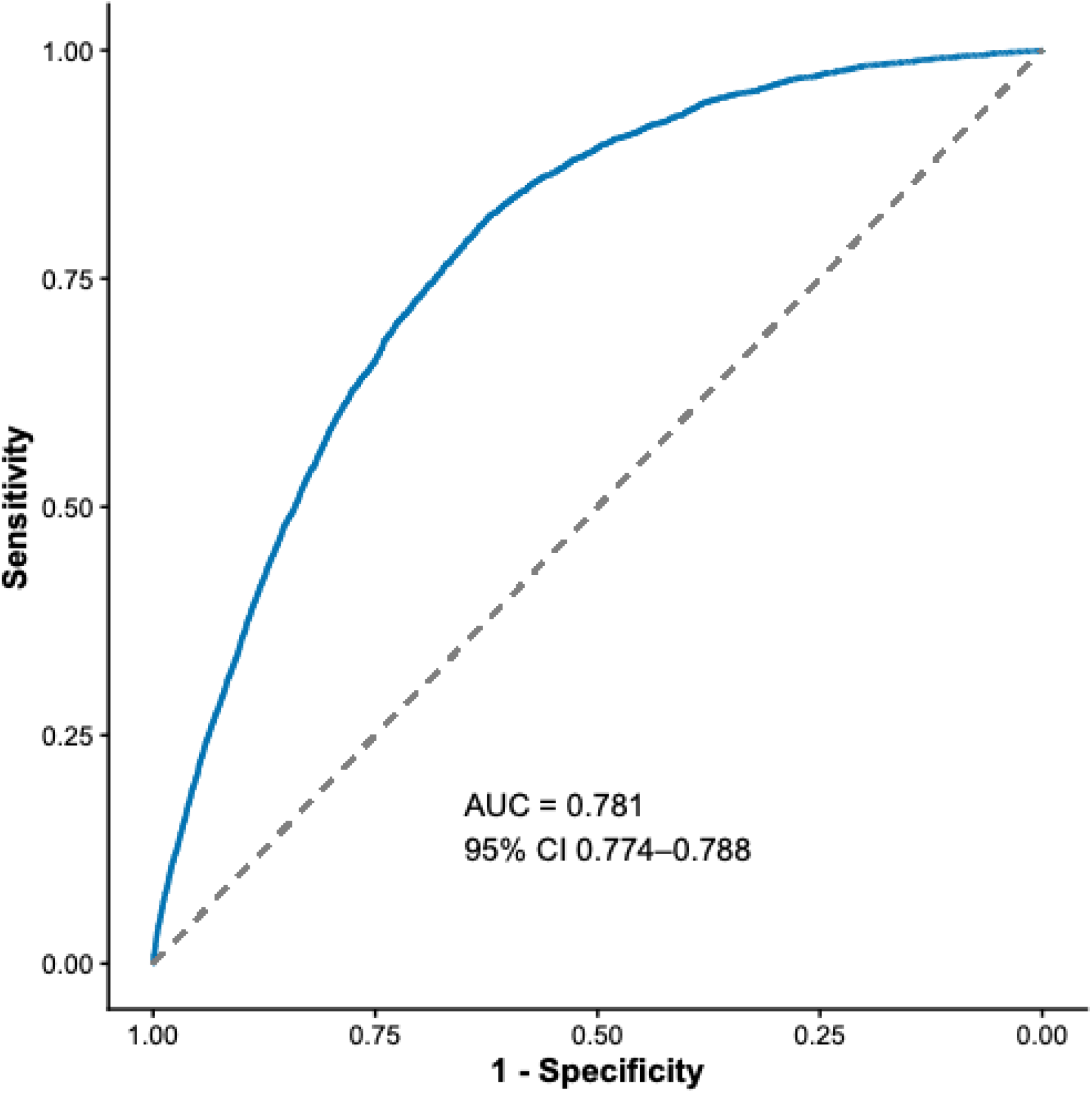
ROC curve showing the specificity and sensitivity of the multivariable logistic regression model

### Forest Plot of Adjusted Odds Ratios

A forest plot summarizing the adjusted odds ratios and corresponding 95% confidence intervals is presented in Figure 3. The strongest positive associations with stroke were observed for poorer general health, diabetes mellitus, current smoking, multiracial ethnicity, and Black race, whereas regular physical activity, higher income, Asian race, and Hispanic ethnicity were independently associated with lower odds of stroke.

**Figure 3:**
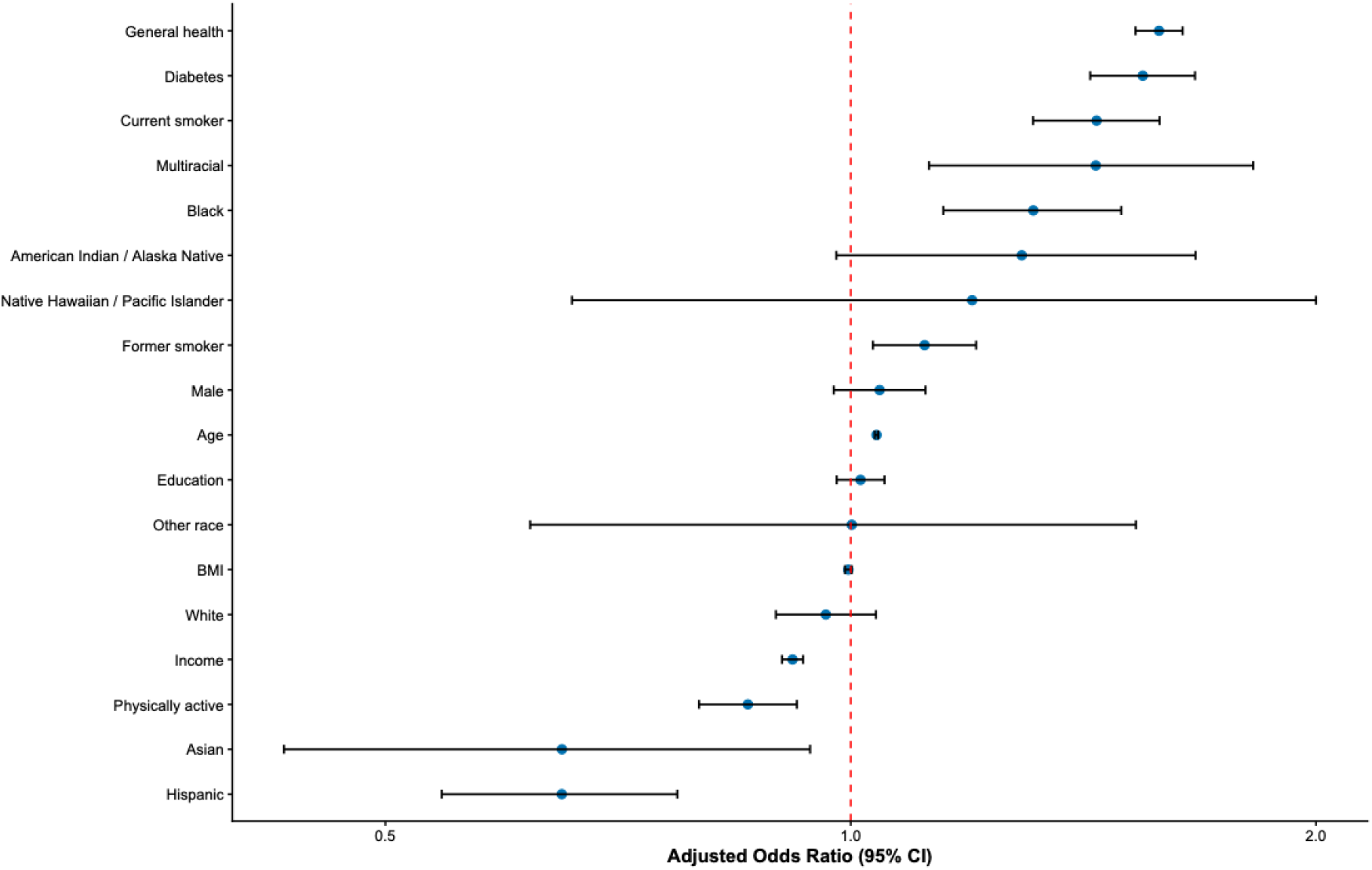
Forest Plot summarizing the adjusted odds ratios (aORs) for various predictive variables

## Discussion

The present study investigated demographic, socioeconomic, lifestyle, and clinical predictors of self-reported physician-diagnosed stroke using pooled nationally representative BRFSS data collected between 2021 and 2023. After adjustment for potential confounders using survey-weighted multivariable logistic regression, increasing age, diabetes mellitus, current smoking, poorer self-rated general health, Black race, and multiracial ethnicity were independently associated with higher odds of stroke, whereas regular physical activity, higher household income, Asian race, and Hispanic ethnicity were associated with lower odds. Furthermore, the multivariable model demonstrated good discriminative performance (AUC = 0.781), indicating that routinely collected demographic, socioeconomic, and behavioral variables can effectively identify individuals at increased risk of stroke in population-based settings. These findings are consistent with the growing body of epidemiological evidence showing that the majority of stroke risk is attributable to a relatively small number of modifiable vascular and lifestyle-related factors (1,11–13).

Age emerged as one of the strongest independent predictors of stroke, with each additional year associated with an approximate 4% increase in stroke odds. This observation is consistent with the Global Burden of Disease Study and the Framingham Stroke Risk Profile, both of which identify advancing age as the most important non-modifiable determinant of stroke risk (1–3). Age-related vascular remodeling, endothelial dysfunction, arterial stiffness, chronic inflammation, and cumulative exposure to cardiovascular risk factors progressively increase susceptibility to cerebrovascular disease. Recent meta-analyses have similarly demonstrated that stroke incidence increases exponentially after the sixth decade of life, emphasizing the need for early identification and preventive interventions before advanced age is reached (8,11).

Diabetes mellitus remained a major independent predictor of stroke, with diabetic individuals demonstrating approximately 55% higher odds of stroke after adjustment for potential confounders. Diabetes accelerates atherosclerosis through endothelial dysfunction, oxidative stress, chronic inflammation, platelet activation, and microvascular injury, thereby increasing the risk of both ischemic and hemorrhagic stroke. These findings are consistent with the INTERSTROKE study, which identified diabetes as one of the leading modifiable vascular risk factors worldwide (12). More recent systematic reviews and meta-analyses have confirmed that diabetes substantially increases stroke risk across diverse populations and emphasize that optimal glycemic control remains fundamental for reducing cerebrovascular complications (8,9).

Smoking also demonstrated a strong independent association with stroke. Current smokers exhibited markedly greater odds of stroke than never smokers, while former smokers continued to show elevated risk, although to a lesser extent. Cigarette smoking promotes vascular injury through oxidative stress, endothelial dysfunction, increased platelet aggregation, inflammation, and accelerated atherosclerosis. Although smoking cessation substantially reduces vascular risk over time, residual risk among former smokers likely reflects cumulative vascular damage before cessation. These findings are in agreement with the INTERSTROKE investigation and contemporary systematic reviews identifying tobacco use as one of the most preventable causes of stroke worldwide (8,9,12).

Regular physical activity was independently associated with significantly lower odds of stroke, supporting extensive evidence demonstrating the cardiovascular benefits of regular exercise. Physical activity improves endothelial function, reduces blood pressure, enhances insulin sensitivity, decreases systemic inflammation, and contributes to healthier body weight, thereby reducing overall vascular risk. Recent systematic reviews have consistently demonstrated that individuals engaging in moderate-to-vigorous physical activity experience substantially lower stroke incidence than sedentary individuals, reinforcing current public health recommendations promoting regular physical activity as a cornerstone of primary stroke prevention (8,9).

An important finding of the present study was the strong association between self-rated general health and stroke. Participants reporting poorer general health exhibited significantly higher odds of stroke after adjustment for conventional vascular risk factors. Self-rated health integrates multiple dimensions of health status, including chronic disease burden, physical functioning, mental well-being, frailty, and healthcare utilization. Previous epidemiological studies have demonstrated that self-perceived health independently predicts cardiovascular disease, hospitalization, disability, and mortality, suggesting that this simple patient-reported measure captures aspects of overall health that may not be fully reflected by individual clinical variables.

Socioeconomic disparities also played a significant role in stroke risk. Higher household income was independently associated with lower odds of stroke, whereas educational attainment was not independently associated after multivariable adjustment. Income likely reflects current access to healthcare, affordability of preventive services, nutrition, housing conditions, and the ability to maintain healthier lifestyles. A recent systematic review of socioeconomic inequalities in stroke concluded that lower socioeconomic position contributes to increased stroke incidence through multiple interconnected pathways, including healthcare access, behavioral risk factors, psychosocial stress, environmental exposures, and reduced opportunities for disease prevention (10). These findings emphasize that reducing stroke burden requires not only clinical management of vascular risk factors but also broader public health strategies addressing social determinants of health.

The racial and ethnic disparities observed in this study are also consistent with previous epidemiological evidence. Black participants demonstrated significantly higher odds of stroke, whereas Asian and Hispanic participants exhibited lower odds after adjustment for demographic, socioeconomic, and behavioral characteristics. These differences likely reflect a complex interaction among genetic susceptibility, environmental exposures, socioeconomic inequalities, healthcare accessibility, and differences in the prevalence and management of vascular risk factors. Similar disparities have been reported in national surveillance data and international epidemiological studies, highlighting persistent inequalities in stroke burden across racial and ethnic populations (1,11–13). The elevated odds observed among multiracial individuals warrant further investigation because this population remains underrepresented in stroke research.

The multivariable logistic regression model demonstrated good discriminative performance with an AUC of 0.781, indicating satisfactory ability to distinguish individuals with and without stroke using routinely collected public health variables. Although the model is not intended to replace comprehensive clinical risk prediction tools, it demonstrates that demographic, socioeconomic, behavioral, and self-reported clinical information can effectively support population-level risk stratification. Such models may be particularly valuable for surveillance, community screening, and public health planning in settings where detailed clinical investigations are unavailable or impractical.

The present study has several notable strengths. First, the analysis utilized a large nationally representative sample comprising more than 235,000 U.S. adults from three consecutive BRFSS survey cycles, thereby providing substantial statistical power and broad generalizability. Second, survey-weighted regression analysis appropriately accounted for the complex sampling design of the BRFSS, generating nationally representative effect estimates. Third, multiple demographic, socioeconomic, behavioral, and clinical predictors were evaluated simultaneously, allowing estimation of independent associations while minimizing confounding. Finally, the study followed established recommendations for observational epidemiological analyses and prediction modelling, including appropriate consideration of study design, multivariable adjustment, and assessment of model discrimination (5–7,14,15).

Several limitations should be acknowledged. First, the cross-sectional design precludes causal inference, and temporal relationships between risk factors and stroke cannot be established. Second, both stroke status and predictor variables were self-reported, introducing the possibility of recall bias and outcome misclassification. Third, residual confounding from unmeasured variables—including atrial fibrillation, lipid concentrations, medication use, dietary factors, and genetic susceptibility— cannot be excluded. Fourth, because the BRFSS surveys only non-institutionalized adults, individuals residing in hospitals or long-term care facilities were not represented, potentially limiting generalizability to these populations. Finally, although the model demonstrated good discrimination, external validation in independent international cohorts would further establish its applicability across diverse populations.

## Conclusion

In conclusion, this nationally representative survey-weighted analysis identified increasing age, diabetes mellitus, cigarette smoking, poorer self-rated general health, Black race, and multiracial ethnicity as significant independent predictors of stroke among U.S. adults, whereas regular physical activity, higher household income, Asian race, and Hispanic ethnicity were associated with reduced stroke risk. The multivariable model demonstrated good discriminative performance, highlighting the utility of routinely collected demographic, socioeconomic, and behavioral information for population-level stroke risk assessment. These findings reinforce the importance of integrated public health strategies that combine lifestyle modification, chronic disease prevention, equitable healthcare access, and interventions addressing social determinants of health to reduce the burden of stroke.

## Data Availability

Data is publicaly available through CDC BRFSS

